# The Politics of Vaccine Hesitancy in the United States

**DOI:** 10.1101/2021.12.01.21267160

**Authors:** Jian Cao, Christina Ramirez, R. Michael Alvarez

## Abstract

**Objective:** Why are Americans COVID-19 vaccine hesitant? We test social science hypotheses for vaccine hesitancy, focusing on partisanship, trust in institutions, and social-demographic characteristics of registered voters.

**Methods:** We use survey data from a representative sample of American registered voters collected in November 2020 to study vaccine hesitancy, and the reasons for vaccine hesitancy, at a point in time before the vaccine was available and hence show underlying responses based on beliefs and not on clinical trial data. We use multivariate logistic regression models to test hypotheses on vaccine hesitancy.

**Results:** We find that consistently similar groups of people tend to be vaccine hesitant. Specifically, Black voters, those between the ages of 45 and 64, female voters, voters without college degrees, voters not worried about the spread of COVID-19, and voters who are concerned about government and the CDC’s handling of the COVID-19 pandemic, were vaccine hesitant. We also provide intriguing results showing the nuanced reasons that the vaccine hesitant provide.

**Conclusions:** Our analysis allows us to establish important baseline information from a social science perspective on vaccine hesitancy at a crucial time, right before COVID-19 vaccines were beginning to be made available to adult Americans. What emerges from our analysis is a nuanced perspective on vaccine hesitancy in the United States, from this important point in the history of the COVID-19 pandemic.

## 1 Introduction

On January 21, 2020, the United State’s Centers for Disease Control (CDC) announced confirmation of the first 2019 novel coronavirus (COVID-19) case in the United States. Within ten days, the World Health Organization (WHO) declared a global health emergency due to COVID-19, and on February 3, 2020 the U.S. government declared a public health emergency. And just a month later, on March 11, 2020, WHO declared that COVID-19 was a pandemic. In recognition of the need for rapid development of effective vaccines to protect U.S. residents against COVID-19, the U.S. government launched “Operation Warp Speed” (OWS) in May 2020, with an initial target of 300 million doses of COVID-19 vaccine available by January 2021 (GAO, 2021). Vaccinations in the U.S. began in late December 2020, and currently three different vaccines are available in the U.S. (Pfizer-BioNTech, Moderna, and Johnson & Johnson) under Emergency Use Authorization (EUA).^1^ As of November 4, 2021, Center for Disease Control (CDC data) show that 222 million U.S. residents have received at least one dose of available vaccine (about 67% of the total population), while over 192 million U.S. residents are considered fully vaccinated (58% of the total population).^2^

While COVID-19 vaccinations in the U.S. accelerated in the winter and spring of 2021, there are clear signs currently that the number and rate of U.S. vaccinations have been increasing at a much slower rate in the summer and fall of 2021. This has raised concerns about COVID-19 vaccine hesitancy in the United States and globally, as without widespread acceptance of the various vaccines being offered, the population may not see sufficient levels of vaccination to achieve “herd immunity” that will be sufficient to end the pandemic.

This begs a number of important questions — why, despite the initial interest in COVID-19 vaccinations, has demand for them waned? Was this initial surge in demand for vaccinations, which peaked in less than four months, predictable given pre-existing concerns about vaccinations in general, and the overall politicization of COVID-19 vaccinations in the U.S.? And finally, is there anything that can be done to change these current trends — what can be done to persuade those who are vaccine hesitant in the U.S. to get the shots? Shedding light on these important questions is the goal of our paper.

In the United States, Pfizer’s vaccine was the first to receive Emergency Use Authorization on December 11, 2021.^3^ Pfizer released the first interim results from their Phase 3 clinical trial in a press release on November 9, 2020,^4^ and published the results on December 31, 2020 (Polack et al., 2020). As much of this clinical trials data was publicly available after our survey data was collected (November 2020), our data reflect pre-existing beliefs about COVID vaccines and vaccines in general. The survey data we use in this paper were gathered from representative voters who most likely did not have early access to vaccine clinical trial data and thus their attitudes towards COVID vaccines most likely reflect partisanship attitudes and pre-existing beliefs, which have been shown to be a strong predictor of COVID-19 policy preferences and behaviors (Grossman et al., 2020). That the data we use in this paper come from an important moment in the distribution of COVID vaccines in the United States is an important contribution of our work.

In this paper, we examine the issue of COVID-19 vaccine acceptance from a social science perspective before any data on the vaccine was widely available. Thus, the opinions formed on whether one would accept a vaccine or not are only based on pre-existing beliefs and not facts, and should be less affected by misinformation about COVID vaccines. Using a large and representative sample of registered voters interviewed immediately after the November 2020 election, we study vaccine acceptance from different behavioral, attitudinal, and political perspectives. Our study highlights a central difficulty regarding COVID-19 vaccine acceptance in the United States: the reasons for vaccine hesitancy are complex and nuanced among Americans. It’s not a simple matter of partisan polarization, or the resistance in that might be amenable to simple types of persuasion. While vaccine hesitancy in the United States is associated with partisanship, it’s also associated with where adults get their news, their trust in different institutions, and their social-demographic identities. In short, it’s complicated – meaning that there is no simple and easy way to quickly increase vaccination rates in the United States.

## 2 What Determines Vaccine Hesitancy?

Vaccine hesitancy as a general issue is common throughout the world. In 2019, the World Health Organization listed vaccine hesitancy as one of the top ten threats to global health.^5^ Some the most common reasons for hesitancy are perceptions of risks and benefits, a lack of knowledge about their use and efficacy, and in many places, religious beliefs (Larson et al., 2014; Lane et al., 2018; Kumar et al., 2016).

In June of 2020, while COVID-19 vaccines were still in human trials, a global survey was conducted with 13,426 respondents in 19 countries to assess the acceptance of a COVID vaccine (Lazarus et al., 2020). While there were important cross-national differences, with acceptance ranging from 90% in China, to 74.5% in the United States and 55% in Russia, overall vaccine acceptance was high with 71.5% of respondents saying they “completely agree” or “somewhat agree” to the statement “If a COVID-19 vaccine is proven safe and effective and is available, I will take it” (with only 14.2% disagreeing somewhat or completely). However vaccine acceptance in this survey dropped to 48.1% when asked whether “You would accept a vaccine if it were recommended by your employer and was approved safe and effective by the government” with almost 26% disagreeing. Importantly, this finding held across nationalities and even in countries with high reported vaccine acceptance. All respondents reported that they would be less likely to accept a vaccine if it were mandated by employers, suggesting that voluntary compliance could lead to more vaccine acceptance versus vaccine mandates. Studies have shown heterogeneity in vaccine acceptance worldwide with variability over time (Dzieciolowska et al., 2021; Almaghaslah et al., 2021; Dzinamarira et al., 2021; Kukreti et al., 2021; Machida et al., 2021; Neumann-Böhme et al., 2020). World-wide vaccine acceptance is critical to the control of the pandemic and illustrates the necessity of a global effort to increase trust in vaccines.

The Kaiser Family Foundation (KFF) found that in December 2020, 71% of Americans surveyed would agree to get a vaccine if it were available for free and deemed safe by scientists, while 27% remained vaccine hesitant. They found that 35% of Blacks are vaccine hesitant with 47% citing that they don’t trust vaccines in general. They also found that 42% of Republicans are vaccine hesitant with 59% citing side effects as a concern and 55% citing lack of trust in the government to ensure the vaccine’s safety and efficacy.

In this same KFF survey, they found that 30% of American healthcare workers serving on the front lines are vaccine hesitant and as of March 7, 2021, 48% of healthcare workers had not yet received their first dose of the vaccine. They found racial disparities among healthcare workers, with only 39% of Black and 44% of Hispanic healthcare workers reporting getting a vaccine compared to 57% of White healthcare workers. They reported that 28% of Black health care workers do not plan on getting vaccinated. The biggest factors among healthcare workers for vaccine hesitancy are concerns about possible side effects (82%), that these vaccines are too new and they want to wait and see (81%), and a lack of trust in the government to make sure the vaccines are safe and effective (65%). A total of 36% of frontline workers say they are not confident that the vaccines have been properly tested for safety and effectiveness. Notably 53% of Black healthcare workers are not confident compared to 47% of Black adults in the general population. They see similar trends for healthcare workers regarding party identification, with 40% of Republicans not confident and 28% of Democrats not confident. A study done by Rand among Black community stakeholders also found vaccine hesitancy among Black healthcare workers with 48% responding they would not get vaccinated (Bogart et al., 2021). Others have found similar reasons for hesitancy among healthcare workers (Dzieciolowska et al., 2021; Schrading et al., 2021). Given that many people get their healthcare information from their health providers, this hesitancy could lead to reduced acceptance among the general population (MacDonald, 2015).

The John Hopkins Center for Communication Programs conducted a survey in 23 countries in 19 waves with the most recent wave between March 15-March 29, 2021, with 3,949 survey respondents.^6^ Currently, 60.7% of the respondents in the two week period at the end of March said they would accept a COVID-19 vaccine. Interestingly in this survey, 51% of respondents said they had exposure to journalists, but only 10% had trust in them. Similarly, only 36% responded that they had trust in government health officials and only 4% of respondents stated that they had trust in politicians. The highest trust levels were reported for scientists with 68% people saying they have trust in scientists.

Of course, there is detailed literature on vaccine hesitancy that we can draw upon to structure the analysis of our data, as it is a global issue (Larson, 2020; Larson et al., 2014; Lane et al., 2018; Kumar et al., 2016). Prior to the COVID-19 pandemic, there were well-publicized accounts of vaccine hesitancy by parents with children on the autism spectrum (Goin-Kochel et al., 2020), boycotts of the polio vaccine (Jegede, 2007; Murakami et al., 2014), resistance regarding the influenza vaccine (Suryadevara et al., 2014), meningococcal C (Timmermans et al., 2005) and H1N1 influenza (Mesch and Schwirian, 2015). There also is a useful comparative literature on pre-COVID-19 vaccine hesitancy (Hornsey, Harris and Fielding, 2018; Lane et al., 2018; Lazarus et al., 2020; Neumann-Böhme et al., 2020; Wagner et al., 2019). And as COVID-19 vaccinations have rolled out throughout the world in recent months, there have been numerous studies of COVID-19 vaccine hesitancy from across the world, for example, Australia (Edwards et al., 2021), Canada (Dzieciolowska et al., 2021), Japan (Machida et al., 2021), the United Kingdom and Ireland (Murphy et al., 2021; Robertson et al., 2021), the United States (Malik et al., 2020; Schrading et al., 2021), South Africa and Zimbabwe (Dzinamarira et al., 2021), and Taiwan (Kukreti et al., 2021).

The research on vaccine hesitancy focuses on a set of different factors that are associated with individual unwillingness to get a vaccine (or to allow their children to obtain one). Thus the past research helps to frame hypotheses we will test in this paper.

First, an important factor associated with vaccine hesitancy in the United States is race and ethnicity (Bogart et al., 2021; Schneider et al., 2001; Timmermans et al., 2005). In particular, the Black community has a history of mistrust of institutions and of medical researchers specifically. This institutional distrust can be seen clearly in the wake of recent events like the murder of George Floyd, and the subsequent Black Lives Matter protests. Distrust of health care institutions and medical researchers among Blacks in the U.S. has its origins in their experience with past research abuses like those suffered by Black participants in the infamous Tuskegee experiments. This leads to our first hypothesis, that relative to other racial and ethnic minorities, **Blacks are more likely to be vaccine hesitant, more skeptical of the COVID-19 vaccine, and uncertain about how it works**.

Additionally, researchers have focused on other demographic factors, in particular age (Adams et al., 2021). There are many reasons to expect that age and COVID-19 vaccine hesitancy might be associated. Younger people may not have easy access to health care, and may be concerned about the possible costs or side effects of the COVID-19 vaccine. They may perceive that because they are younger, and generally healthy, that they do not need the vaccine. Or they may be less informed about vaccinations in general, and COVID-19 vaccines specifically, than older Americans. These lead us to our second hypothesis, **younger Americans are more likely to be vaccine hesitant, to believe that they may not need the COVID-19 vaccine, to be concerned about the expense, and be worried about possible side effects**.

Perceptions of risks may also be important determinants of COVID-19 vaccine hesitancy, of two different sorts. Of course, people may be concerned about the risk of the vaccine itself. But on the other hand, people may be concerned about the risks of not being vaccinated, perhaps based on their or their family’s experiences with COVID-19. Those who have been immediately impacted by COVID-19, who have seen family or friends catch the virus, or who lost their jobs or income due to the virus, might be more willing to get fully vaccinated. Thus, our third hypothesis is that **those who have had recent negative experiences with COVID-19, who perceive that COVID-19 is a problem in their communities, or who have lost jobs or income due to the pandemic, may be less likely to be vaccine hesitant.**

Another related factor is information about the vaccine and vaccination process. There has been a great deal of misinformation spread about COVID-19 vaccines, whether inadvertent or deliberate, and those who are less informed about vaccines and the COVID-19 vaccinations might be more hesitant to get the shot (Geńe et al., 1992; Kata, 2012; Loomba et al., 2021). Also, there is some evidence that right-wing social media and news outlets may be more likely to spread misinformation about COVID-19 vaccines (Baines, Ittefaq and Abwao, 2021) — those who get news and information primarily from right-wing sources could be more likely to be vaccine hesitant. Given this, we hypothesize that **those who are less informed about the vaccines, or who primarily obtain their news and information from right-wing news sources, may be more vaccine hesitant, and more likely to be skeptical of vaccines**.

There are two final factors that we will examine in our analysis. The first of those is trust in institutions. Past research has noted the role of institutional distrust in vaccine hesitancy (Jamison, Quinn and Freimuth, 2019; Ruijs et al., 2013; Suk, Lopalco and Celentano, 2015). Thus, we hypothesize that **those who are more trusting of the institutions that are promoting COVID-19 vaccines, for example the CDC, might be less vaccine hesitant, and less skeptical of vaccines.**

Finally, in the United States, political reactions to the pandemic have often been polarized on partisan lines, for example with Democratic states more likely to use more restrictive measures to prevent the spread of the virus than Republican states. This may also polarize opinions about vaccines along partisan lines (Grossman et al., 2020). Political beliefs and ideology may more generally structure how people perceive vaccines and vaccinations (Baumgaertner B, 2018). Finally, we hypothesize that **Republicans will be more vaccine hesitant than Democrats, and to express skepticism, a lack of understanding about how the vaccines work, and to not believe that they need to be vaccinated**.

## 3 Data and Methods

### 3.1 Data

We use data from a large, nationally-representative survey of U.S. registered voters. The survey was fielded by YouGov, using respondents from their opt-in panel as well as data from another external provider. The survey was fielded November 4-10, 2020, with an array of questions about the November 2020 election, in particular voting experiences; also included were a variety of questions about issues and economic experiences, including opinions about the COVID-19 pandemic, effects of the pandemic on the respondent and their family, and opinions about COVID-19 vaccinations. It is important to note that the Pfizer press release on the preliminary efficacy results was announced on November 9, 2020. While, it is possible that some people in our survey may have heard the announcement on the last day of the survey, it was not approved (or even reviewed by the FDA) as of that date. A total of 5,051 respondents participated in the survey, with an oversampling of respondents from California. We weight the sample based weights provided by YouGov using gender, age, race, education, U.S. Census region, state of residence, and 2020 Presidential vote, in order to obtain a nationally-representative sample of the U.S. electorate. The weights range from 0.1 to 5.973, with a mean of 1 and a standard deviation of 1. The margin of error for the survey is *±* 2.

### 3.2 Asking About Vaccine Hesitancy

Our work in this paper is based on two important questions in our survey. The weighted topline results for these two questions are provided in Table 1.^7^ Our primary question about vaccine acceptance was asked to all of the respondents to our survey, “If a vaccine for COVID-19 were available today would you get the vaccine?” The second question, asked only to those who responded that they would not get a COVID-19 vaccine, asked them to select up to two reasons regarding why they would not get a vaccine.

**Table 1:**
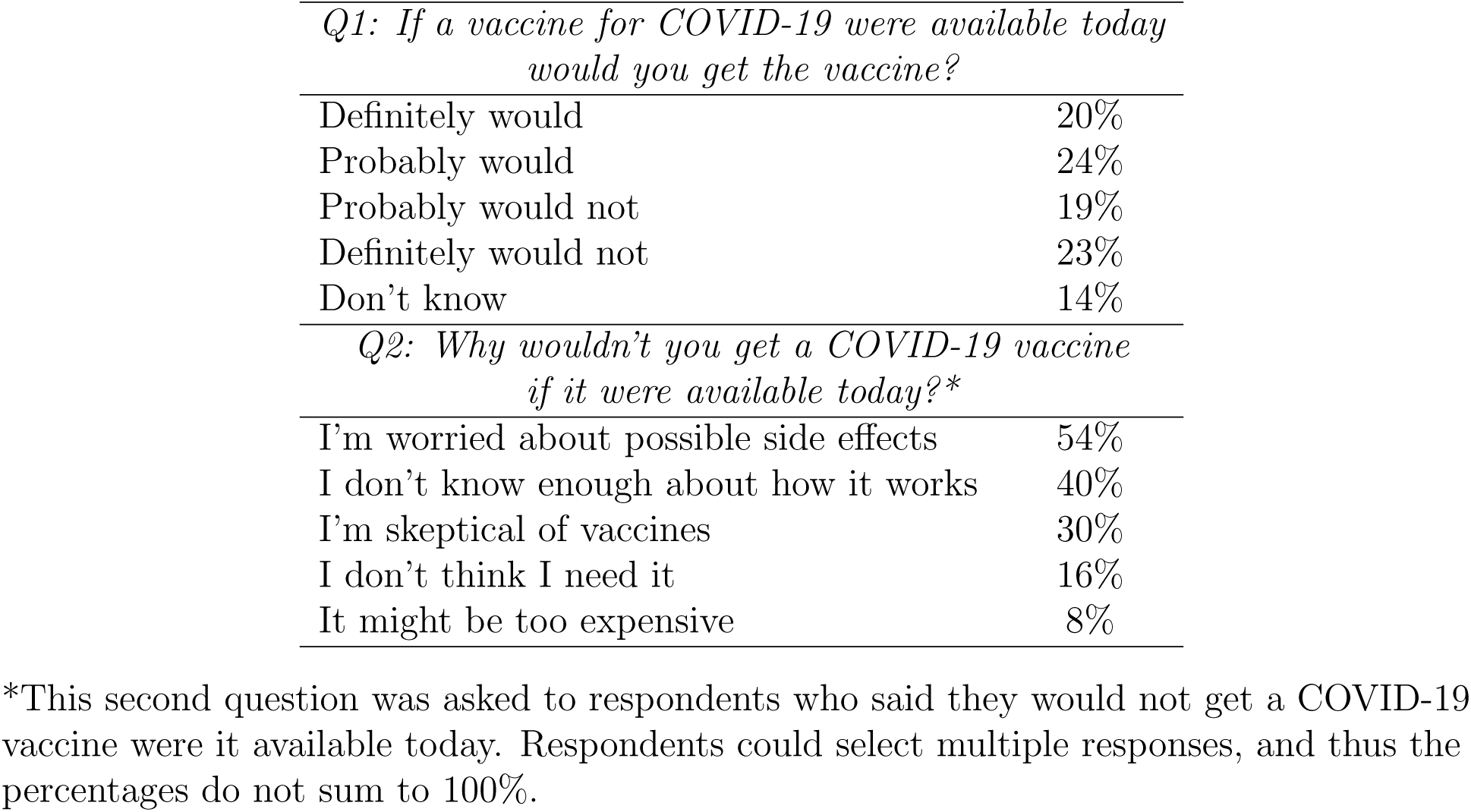
Vaccine Acceptance.

| Table 1: **Vaccine Acceptance.** |  |
| --- | --- |
| *Q1: If a vaccine for COVID-19 were available today would you get the vaccine?* |  |
| Definitely would | 20% |
| Probably would | 24% |
| Probably would not | 19% |
| Definitely would not | 23% |
| Don’t know | 14% |
| *Q2: Why wouldn’t you get a COVID-19 vaccine if it were available today?\** |  |
| I’m worried about possible side effects | 54% |
| I don’t know enough about how it works | 40% |
| I’m skeptical of vaccines | 30% |
| I don’t think I need it | 16% |
| It might be too expensive | 8% |

\*This second question was asked to respondents who said they would not get a COVID-19 vaccine were it available today. Respondents could select multiple responses, and thus the percentages do not sum to 100%.

As shown in Table 1, in our sample 20% said they definitely would get a COVID-19 vaccine, while 24% said they probably would. Thus 44% of our sample would be likely to get a vaccination were it available to them at the time of the survey. We also see that 19% said they probably would not get the vaccine, and that 23% said they definitely would not. In our survey, 14% said they did not know whether they would or would not get the vaccine at the time the survey was conducted.

For the respondents who indicated they would not get a vaccination were it available to them at the time our survey was conducted, we gave them the opportunity to indicate why they would not get vaccinated.^8^ They could mark up to two reasons, from the set of five reasons shown in the lower panel of Table 1 (note that as respondents could mark up to two of these reasons, the column percentages do not sum to 100%).^9^ The most prevalent reason for vaccine refusal was “I’m worried about possible side effects”, which 54% of the respondents indicated. The next most prevalent reason was “I don’t know enough about how it works” (40%) followed by “I’m skeptical of vaccines” (30%).

### 3.3 Multivariate Analysis Methods

In order to delve more deeply into vaccine hesitancy, and to potentially determine why many American registered voters might not get a COVID-19 vaccine were it offered to them, we next turn to a more detailed multivariate analysis of these data. We start with the responses to Q1, “If a vaccine for COVID-19 were available today would you get the vaccine?” While our survey question allows an ordinal response, for easier interpretation we focus here on a binary outcome variable, which is coded as vaccine acceptance (if the respondent said either that they definitely would get the vaccine, or that they probably would) or vaccine hesitancy (if they said that they probably would not get the vaccine, or that they definitely would not). The results from the multinomial model are presented in Table SI6.

We present the details of our multinomial model in the paper’s Supporting Information (Section B). Our outcome variables are the individual’s response to the question of vaccine acceptance with “definitely would” or “probably would” as 1 and “probably would not” or “definitely would not” as 0. The individuals who answered “don’t know” to the vaccine acceptance question are omitted in the binary logistic model (but included in the multinomial logistic model presented in Table SI6). The covariates we use to test our hypotheses include demographic questions such as age, gender, race/ethnicity, region, education, and income; pandemic questions such as attitudes of COVID-19 spread, perceptions of government officials’ performances in handling the pandemic, personal financial situations, loss of income due to the pandemic, and attitudes of the reopen; political questions such as partisanship, political news diet, attitudes of social issues; and behavioral questions such as social media use. We show the results of this binary logistic regression for vaccine acceptance in section 4.1.

Binary logistic regression models are nonlinear, and thus the coefficients from those models are not easy to interpret. Thus, in the Results section, rather than present the logistic regression coefficients, instead we provide transformation of the nonlinear logistic regression coefficients into plots of the average marginal effects of each covariate (and an associated 95% confidence interval). This presentation allows for an easier interpretation of the binary logistic regression results. We present the details about how we estimate the average marginal effect for each covariate (and the confidence interval for the estimate) in the Supporting Information (Section B).

We then provide binary logistic results for the five “excuse” outcomes; conditional on stating that they probably would not or definitely would not get a COVID-19 vaccine, survey respondents could provide up to two of the following reasons:

- I’m worried about possible side effects.
- I don’t know enough about how it works.
- I’m skeptical of vaccines.
- I don’t think I need it.
- It might be too expensive.

Those binary logistic models use the same set of covariates as the models for vaccine acceptance. We also present below the estimated average marginal effects from these logistic regression models, using the same methodology outlined above.

One issue that arises with the use of survey data like ours for multivariate modeling is missing data. Survey respondents can skip and not answer questions, and if we were simply to delete cases with at least one missing covariate we would drop 20.53% of our sample from the analysis, risking biased results. Instead of dropping cases with missing information, we use multiple imputation to produce estimates for cases that lack responses for the covariates we use in our models (specifically we use the implementation of multiple imputation developed by Honaker and King (2010)). Details about multiple imputation are available in the paper’s Supporting Information, Section C.

## 4 Results

In this section, we show the results of the multivariate analyses of the vaccine acceptance and refusal reasons, and examine if the hypotheses in section 2 are supported by evidence from the survey data.

### 4.1 Vaccine Acceptance/Refusal

We first focus on the binary outcome model of vaccine acceptance. In Figure 1, we plot the average marginal effects of the selected covariates as dots and the 95% confidence intervals as bars.^10^ The average marginal effect shows that, if a covariate changes from the reference to the specified state, how much would the probability of vaccine acceptance change. For instance, the average marginal effect of being “very worried” about the COVID spread is 0.252, which means compared to the voters who are “not at all worried”, the “very worried” respondents have a 25.2% higher probability to accept the vaccine. Covariates that have significant (*α <* 0.05) average marginal effects are highlighted in black while the others are in grey.^11^

**Figure 1:**
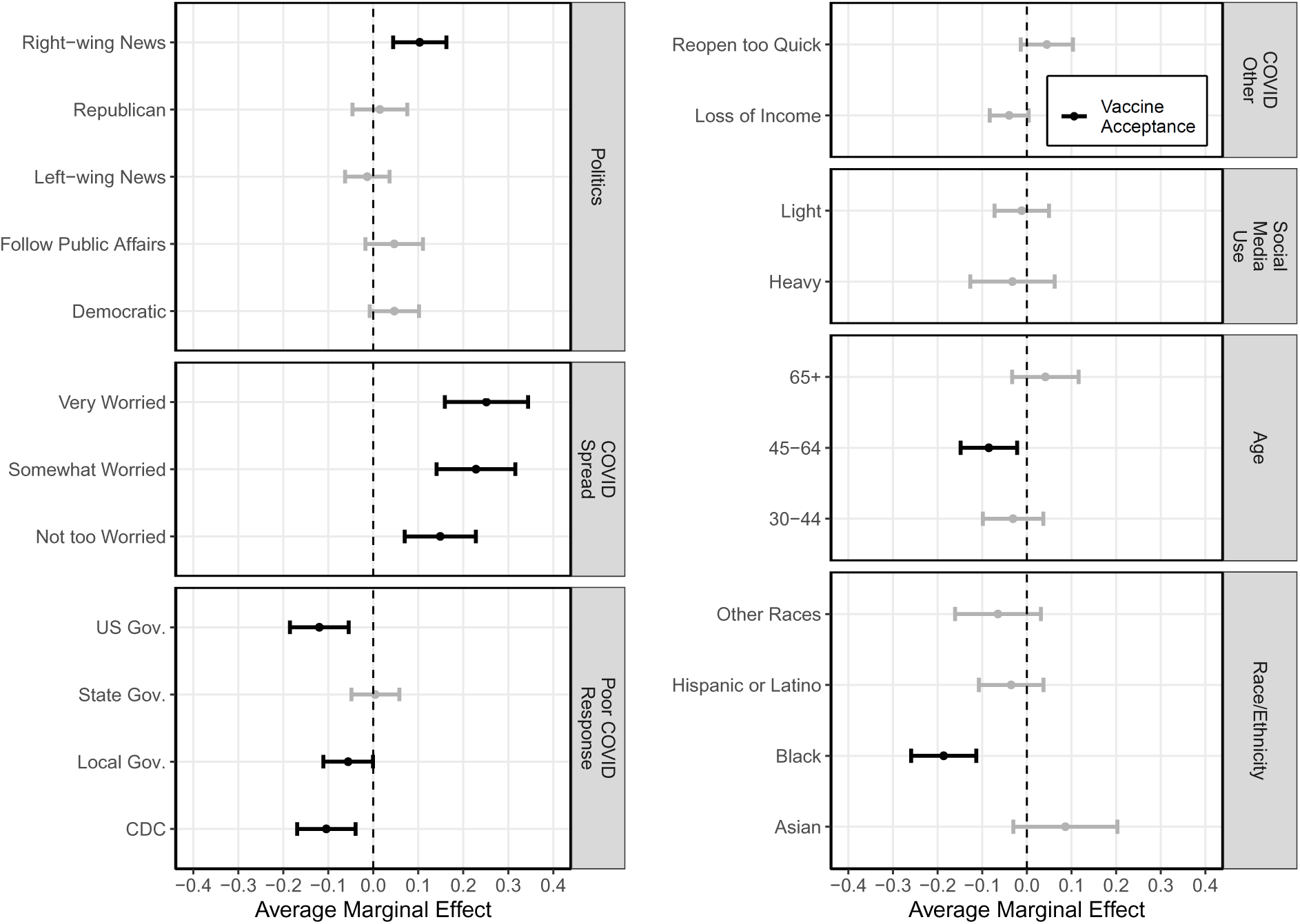
Logit Regression: Vaccine Acceptance

Now we validate our six hypotheses using the evidence from the binary logistic regression. First, compared to White voters, Black voters are less willing to accept the COVID-19 vaccine, which confirms our hypothesis I and is in line with the findings of Kaiser Family Foundation and Bogart et al. (2021). Second, compared to the young voters between 19-29, voters between 45-64 are less willing to accept the vaccine. The other age groups are not significantly different from the young voters in terms of vaccine acceptance. This finding contradicts our hypothesis II. The first half of our hypothesis III is supported by the data, in other words voters who worry about the spread of the COVID-19 pandemic are less likely to be vaccine hesitant, while loss of jobs or income due to the pandemic has no significant effect on vaccine acceptance. For hypothesis IV, we find opposite evidence that voters who primary obtain their news and information from right-wing news outlets^12^ are more willing to accept the vaccine. Next, voters who worry about the federal/local governments’ and CDC’s performances in handling the COVID-19 pandemic are less willing to be vaccinated, which confirms our hypothesis V. Finally, while the literature suggests polarized opinions about vaccines along partisan lines, our data does not support that argument by the date that the survey was fielded.

In addition to the findings related to the six hypotheses, we also find that voters who are male or have college or higher degrees are less likely to be vaccine-hesitant, while voters’ region, income, financial situation, attitudes about COVID-19 reopening, and social media use^13^ do not have significant effects on vaccine acceptance. Moreover, among the covariates, perceptions of COVID-19 spread and race/ethnicity have the largest average marginal effects on vaccine acceptance.

### 4.2 Reasons for Vaccine Refusal

Next, we focus on the voters who “probably would not” or “definitely would not” accept the vaccine and study the reasons behind their decisions. We conduct binary logistic regressions for each refusal reason using the same covariates *X_i_* used for the vaccine acceptance model. The dependent variable *Y ^j^* equals one if voter *i* marked refusal reason *j*, zero otherwise. The study is restricted to the voters who would not accept the vaccine, which is *i ∈ {Y_i_* = 0*}*. The average marginal effects and error bars of the binary logistic regressions for the five refusal reasons are shown in Figure 2. We use shapes to distinguish the refusal reasons: “side effects” — diamonds, “don’t know how it works” — squares, “skeptical of vaccines” — triangles, “don’t need it” — circles, and “too expensive” — upside-down triangles.

**Figure 2:**
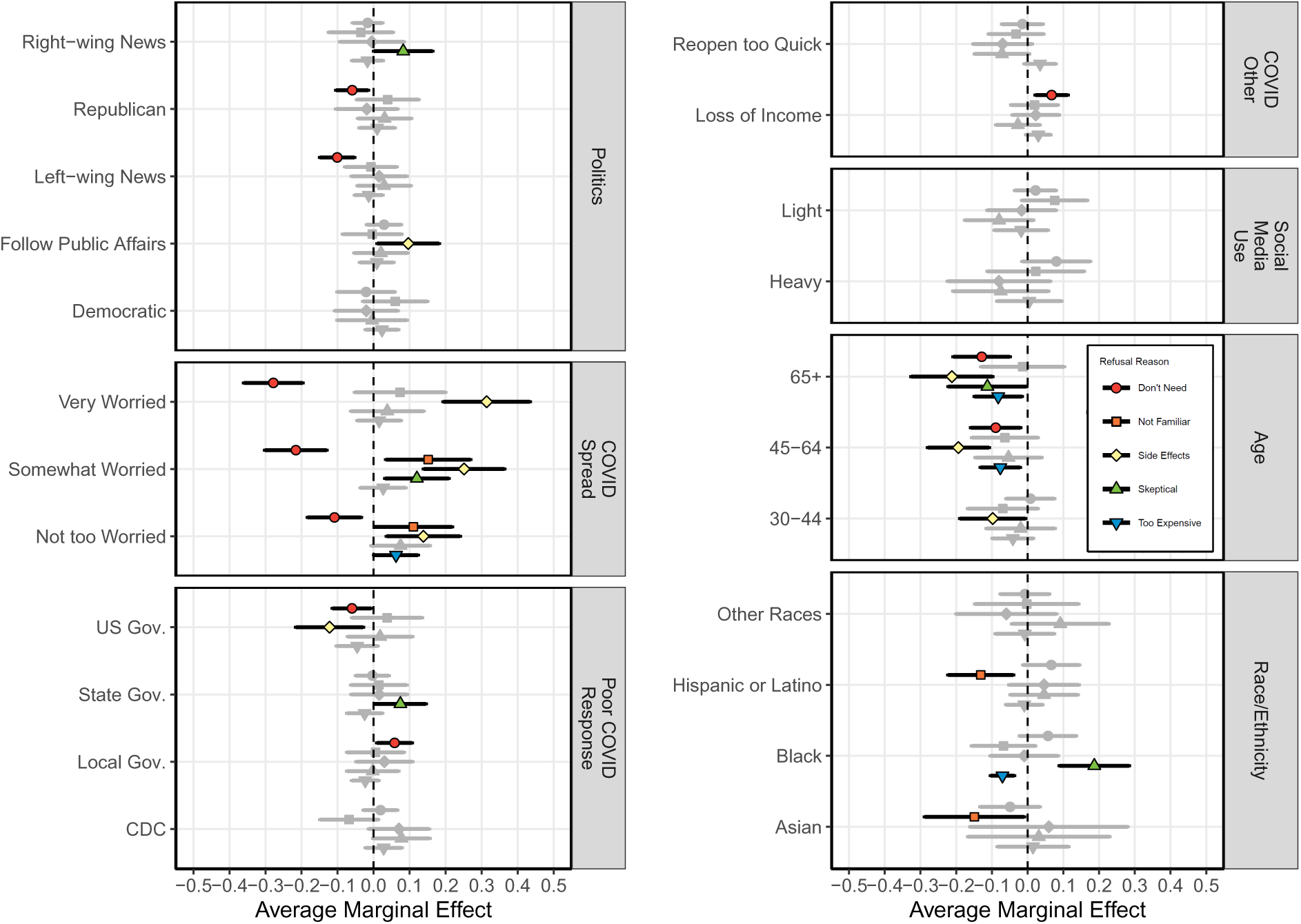
Logit Regression: Refusal Reasons

Here we examine if the results in Figure 2 support our hypotheses. First, Black voters tend to be more skeptical of vaccines, which confirms our hypothesis I. Second, compared to voters above 45, young voters (under 30) worry more about the side effects and expenses and are more likely to believe they may not need the vaccine, which supports the second half of our hypothesis II. Third, voters who mainly follow right-wing news are more skeptical of vaccines; those who mainly follow left-wing news are more likely to believe that they need to be vaccinated. Fourth, those who believe their state governments have done poor jobs in handling the COVID-19 pandemic tend to be skeptical of vaccines. Finally, Republican voters are less likely to believe they do not need the vaccine, but they are not significantly different from the Democratic voters in terms of the likelihood of expressing skepticism and lack of understanding about how vaccines work.

We can also derive the following implications from Figure 2. Voters who often follow public affairs and are concerned about the spread of COVID-19tend to worry more about the vaccine’s side effects. Female voters tend to be more skeptical of vaccines but worry less about the expense of the vaccines. Hispanic and Asian American voters are less likely to express that they are lacking knowledge of the use and efficacy of vaccines. Voters from the south and west tend to mark “Too expensive” more often. And those who attended some college or have college degrees tend to mention side effects as one of their refusal reasons.

## 5 Discussion

In this study we examine data from a large and nationally representative survey of American registered voters regarding COVID-19 vaccine hesitancy, at a time before Pfizer announced the efficacy of their vaccine. Prior to the announcement the FDA stated that they would approve a vaccine with 50% efficacy, thus anchoring people on the 50% number. Thus these results, for the most part, examine the underlying vaccine hesitance with respect to COVID-19 as the real-world efficacy and side effects were not known and not knowable at the time of the survey. Nor should our survey estimates be affected by misinformation about COVID vaccines that may have been disseminated after the Pfizer announcement, and after Americans began to be vaccinated in large numbers. These results may differ from other vaccines such as influenza and common childhood vaccines in that COVID-19 has been very politicized. The topline vaccine hesitancy of this survey is similar to another online survey taken November 18-20, 2020 by the Pew Research Center (Funk, Tyson and Nolan, 2020). They found that 18% of people surveyed would ”definitely NOT get the vaccine” and a total of 39% of people were vaccine hesitant. This compares favorably with our topline numbers of 41.7% of vaccine hesitancy in our survey.

Notably, other surveys have found that even those who are hesitant regarding the COVID-19 vaccine have accepted previous vaccines before, suggesting perhaps a political effect of COVID-19 or skepticism surrounding this vaccine in particular. The YouGov survey offers a rare opportunity to glean insights into the underlying mindset that influence vaccine uptake and hesitancy. Understanding the underpinnings can yield insights into how to increase vaccine uptake in people who a priori are vaccine skeptical. This analysis gives a glimpse into underlying beliefs in the absence of real-world COVID-vaccine data. Strengths of the analysis include using data from registered voters which offers a glimpse into the underlying beliefs about a COVID vaccine that were based, not on data, but on pre-existing beliefs which could have been politicized. Subsequent surveys such as the Kaiser Family Foundation show that those who state they would definitely not get the vaccine remain relatively unchanged despite data being presented with real-world efficacy superior to what had been promised. This suggests that the politicization of vaccine hesitancy is not easily combated by providing positive and persuasive data or information. Perhaps due to high levels of distrust regarding politicians, reporters, and government public health officials as seen in the Johns Hopkins survey, people are skeptical of the data and information provided about vaccine effectiveness. But in that survey, as scientists were identified as trustworthy, perhaps messaging from the scientific community might help restore trust and reduce vaccine skepticism and hesitancy.

Furthermore, we go beyond the simple examination of survey toplines and cross-tabulations, using multivariate choice models that give us the opportunity to control for a wide array of beliefs, opinions, and demographic identities while testing specific hypotheses. We find that consistently similar groups of people, especially in the United States, tend to be vaccine hesitant. Specifically, Black voters, those between the ages of 45 and 64, female voters, voters without college degrees, voters not worried about the spread of COVID-19, and voters who are concerned about government and the CDC’s handling of the COVID-19 pandemic, are vaccine hesitant.

We also provide intriguing results showing the reasons that the vaccine hesitant provide, and find that young voters (under 30) are more likely to be concerned about the side effects, cost, and necessity of the COVID-19 vaccine. Black voters tend to be skeptical of vaccines. Voters with higher education or those worried about the spread of COVID-19 mention side effects more often as reasons to not get vaccinated. Registered Republican voters and those who have left-oriented media diet are less likely to believe they do not need the vaccine. Thus, what emerges is a very nuanced perspective on vaccine hesitancy in the United States, from this important point in the history of the COVID-19 pandemic.

In other surveys, a surprisingly large proportion of healthcare workers remain vaccine hesitant, with many of the groups wishing to see more data and expressing concerns regarding side effects. Knowing which groups and demographic factors are most likely to be vaccine hesitant can guide policy targeting these groups with more information to alleviate their concerns. This is especially important among healthcare providers who often serve as role models for the communities that they serve. But knowing the other segments that are vaccine hesitant, and the reasons behind that hesitancy, can help improve current COVID-19 vaccination rates in the United States in the near future, and also can help health policy makers develop strategies and plans for reducing vaccine hesitance in the future pandemics. Politicization of science is never a good thing. This study shows that vaccine hesitancy is nuanced and complex. There are no one-size-fits-all solutions. To combat vaccine hesitancy, we must truly understand the myriad of reasons behind it, and restore trust in science and public health.

## Data Availability

Data and code will be available upon publication.

## Supporting Information

### A Ethics Statement

The data used in this analysis were collected as part of ongoing research regarding electoral administration and electoral behavior in the United States. The research protocol was reviewed by the Institutional Review Board at the California Institute of Technology, Protocol Number 19-0929, and was determined to be exempt. The survey data were collected by YouGov, primarily using respondents from their opt-in panel, as well as a small sample of panelists from Dynata. The data were collected to study political and social opinions and behaviors at the time of the 2020 presidential election, in both California and the nation. Thus YouGov conducted a national survey (states other than California), along with a California-only survey; for national analyses, like the one reported in this paper, the California oversample (2,532 interviews, 1,891 using YouGov panelists, 641 Dynata panelists) is pooled with the national sample (2,519 interviews, all YouGov panelists) and the responses from California subjects are downweighted. The survey data were provided to us anonymized, with no identifying information included in the data.

Survey respondents self-categorized themselves with respect to the demographic attributes that we study in our analysis, in particular age, race/ethnicity, educational attainment, and gender. The survey questions presented to respondents regarding their demographic identifications are the standard ones used in the survey and polling industry, following industry best practices.

Upon publication, the data and code necessary to replicate the analyses reported in this paper will be available in a public GitHub repository.

### B Methodological Details

The model of interest is defined as:

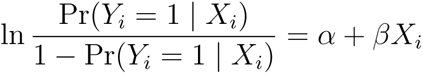

where *Y_i_* is the individual *i*’s response to the question of vaccine acceptance with “definitely would” or “probably would” as 1 and “probably would not” or “definitely would not” as 0. The individuals who answered “don’t know” to the vaccine acceptance question are omitted in the binary logistic model (but included in the multinomial logistic model presented in Table SI6). The covariates *X_i_* include demographic questions such as age, gender, race/ethnicity, region, education, and income; pandemic questions such as attitudes of COVID-19 spread, perceptions of government officials’ performances in handling the pandemic, personal financial situations, loss of income due to the pandemic, and attitudes of the reopen; political questions such as partisanship, political news diet, attitudes of social issues; and behavioral questions such as social media use. We show the results of this binary logistic regression for vaccine acceptance in section 4.1.

We calculate the marginal effect of each covariate *X_k_* for each individual *i*, then calculate the average. The marginal effect of discrete change in categorical variable *k* for individual *i* is obtained as follows:

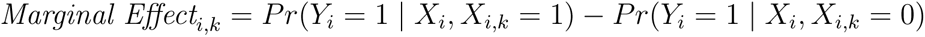

The average marginal effect is then:

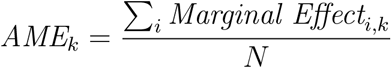

where N is the number of respondents. The confidence interval of *AME_k_* is:

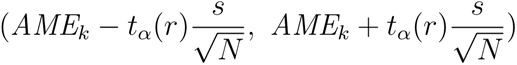

where *t_α_*(*r*) is the critical value from t-distribution. We use a 95% confidence level (*α* = 0.025) and degree of freedom *r* = *N −* 1. 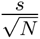 is standard error of *AME_k_*.

### C Multiple Imputation for Survey Data

In this Appendix section we examine the implications of missing values in our survey data. We denote the survey dataset as *Y* , and we use three methods for dealing with the missing data in *Y* : Listwise Deletion (LD), Hot Deck imputation (HD), and Multiple Imputation (MI). Each method produces complete data sets *Y^LD^*, *Y^HD^*, and *Y^MI^* , respectively. The LD method excludes the observations with missing values. In our survey, there are 803 observations that have at least one missing value in the variables included in our main models. After listwise deletion, the resulting data set *Y ^LD^* contains the rest 3,559 complete records. We use Hot Deck (Cranmer and Gill, 2013) method to find the closest alternatives and impute the missing values in target observations, and yield *Y ^HD^* with all 4,362 rows. We also implemented Bootstrapping based Expectation and Maximization (EMB) (Honaker and King, 2010) algorithm to obtain *M* = 5 imputed data sets, 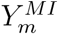 (*m ∈ {*1, 2, 3, 4, 5*}*).

The comparison of average vaccine acceptance computed from *Y^LD^*, *Y^HD^*, and *Y^MI^* are shown in Figure SI1. The listwise deleted data set tend to have higher acceptance rates (i.e. more respondents selected either ”Definitely Would” or ”Probably Would”), but the differences between the methods are not significant (*α* = 0.05).

We repeat the logit regression that with binary outcome acceptance/refusal on data sets *Y^LD^* and *Y^HD^*, and show the average marginal effects in Table SI1. We can see that the average marginal effects that used the listwise-deleted data set differ a lot from those used Hot-Deck imputed and multiple imputed data sets. It is because omitting 18.4% (803 out of 4,362) observations causes a severe information loss, and both estimates of coefficients and standard errors are heavily biased. In addition, although the average marginal effects obtained from multiple imputed data sets are close to those from Hot Deck imputed data sets, the former ones tend to be less significant. It is because Hot Deck imputation couldn’t fully capture the uncertainty of missing values and underestimated the standard errors.

**Figure SI1:**
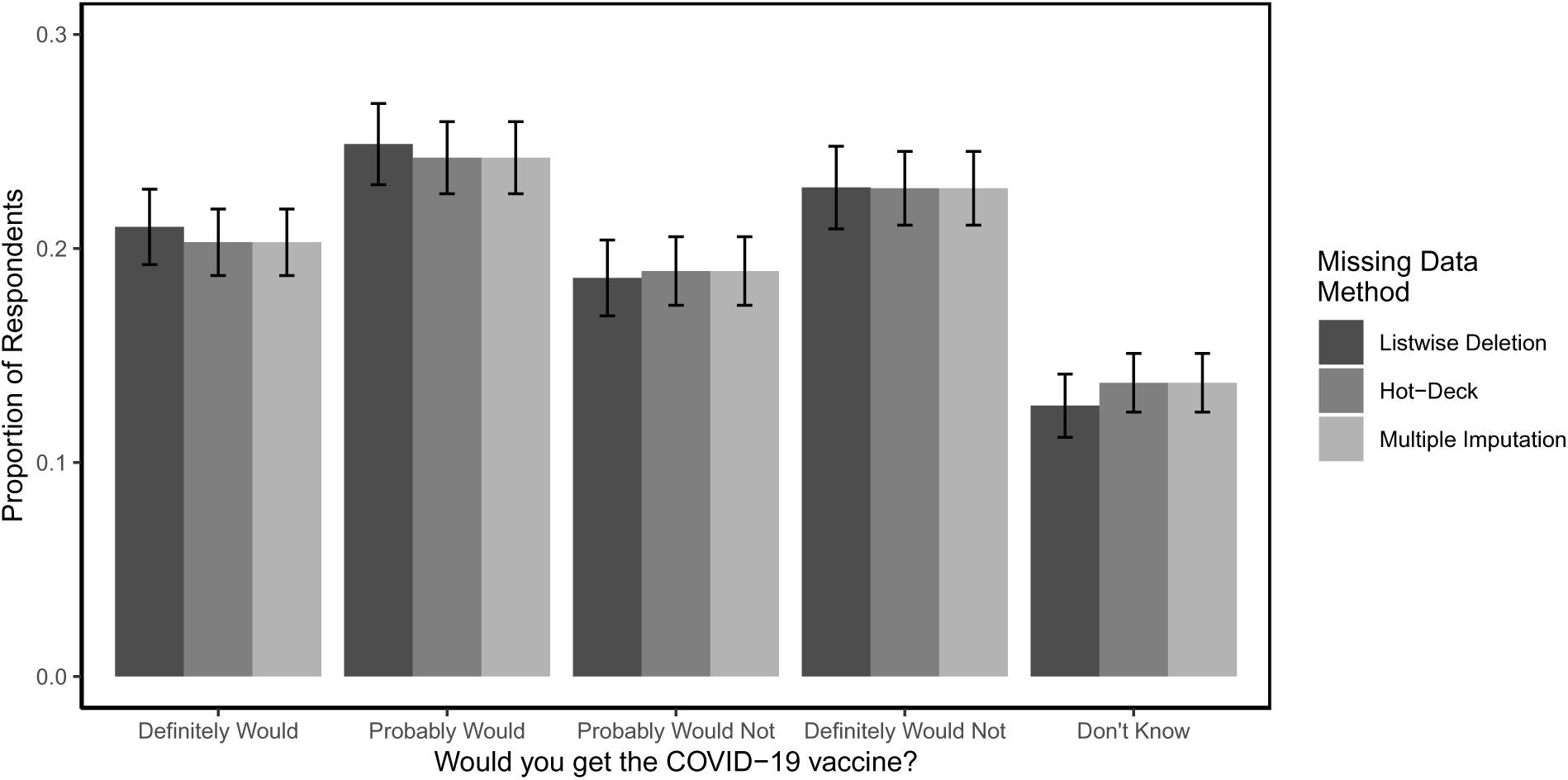
Comparison of Average Vaccine Acceptance over Missing Data Methods

**Table SI1:**
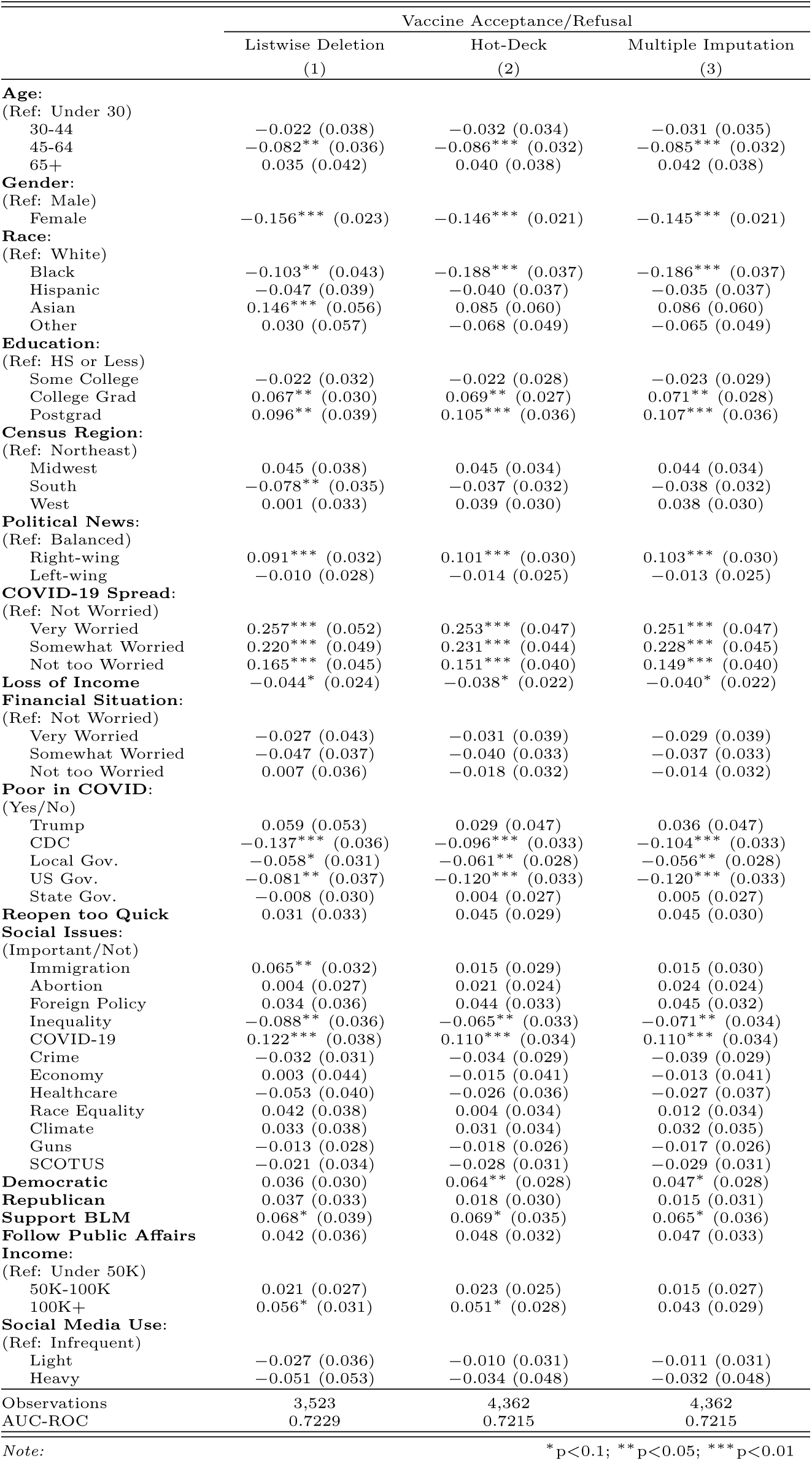
Comparison of Missing Data Methods (Average Marginal Effects)

#### D Bivariate Distributions

In Table SI2 we provide cross-tabulations of responses to our vaccine hesitancy question by selected variables from our survey. The entries in the table are weighted, and are column percentages. Beginning with age, we see something of an age gap in vaccine hesitancy — older registered voters in our survey (those 45 to 64, and 65 and older) were more likely to say they definitely or probably would get vaccinated than were those 30-44 and under 30. We also see a gender gap in vaccine hesitancy in the cross-tabulations: 57% of those who said they would definitely get vaccinated were men, while 52% of those who said that they probably would get the vaccine were men. Sixty-one percent of those who said they definitely would not get the vaccine were women, and 62% of those who said they didn’t know if they would get the vaccine were women.

In our survey cross-tabulations, we see also significant racial and ethnic divides regarding vaccine hesitancy. Blacks and Hispanics are much more likely to be report hesitancy (or that they don’t know if they would get a vaccination) than White and Asian registered voters in our samples. The cross-tabulations provided in Table SI2 also show important differences in vaccine hesitancy by educational attainment levels, in particular on the hesitancy side as much greater proportions of those who probably will not, definitely will not, and who say they don’t know have a high school education or less, or some college. There is also some evidence in Table SI2 to indicate that vaccine hesitancy has a regional component in the United States, as 42% of the probably will not and definitely will not are from the American South.

In the vaccine hesitancy cross-tabulations we also include results for partisanship. The results for these political indicators may not be surprising, given how the rhetoric about COVID-19 and vaccinations was polarized along partisan lines in 2020. Among those who said they would definitely or probably get a vaccination, higher proportions tend to be Democrats. But among those who said they would probably not or would definitely not get the vaccine, we see greater proportions of Republican and Independent voters.

The next set of cross-tabulations are given in Table SI3 (with row percentages given in Table SI5). This table is organized similarly to the previous cross-tabulation, except here the columns provide the weighted proportions of registered voters in our survey who expressed vaccine hesitancy, and who indicated any of the five different reasons for their hesitancy. Examination of Table SI3 by column provides an interesting perspective into the potential groups who might be vaccine hesitant for different reasons. The first reason for hesitancy in our survey was uncertainty regarding potential side effects: looking down the “yes” column for that reason, we see that higher proportions of those aged 45-64, women, Blacks, those with a high school degree or some college, from the South, and Democrats were most likely to provide that reason.

The next reason for vaccine hesitancy in our survey was concerns that it might be expensive. Here we see that most registered voters under 65, Hispanics, those with a high school education or some college, who were from the South, and Democrats indicated cost as one reason for their hesitancy. Next, survey respondents could also indicate that they didn’t think that they needed a COVID-19 vaccine; here we see middle-aged registered voters, men, Blacks, those with a high school education or some college, Southerners, and Republicans providing that reason.

Perhaps most interesting are the cross-tabulations for the two more skeptical reasons for vaccine hesitancy — that the vaccine won’t work or that they are skeptical of vaccines in general. For the former, we see that registered voters 45-64, females, Blacks, those with high school education or some college, from the South, and Democrats expressing that as a reason for their hesitancy. Finally, those who state that they are skeptical of vaccines more generally are different, as they tend to be 45-64 years of age, females, those with a high school education or less, from the South, or Republicans.

##### D.1 Column Percentage Cross-tabulations

**Table SI2:** Vaccine Hesitancy and Selected Variables

|  | Definitely | Probably | Probably Not | Definitely Not | Don't Know |
| --- | --- | --- | --- | --- | --- |
| Total survey | 20.3 | 24.2 | 18.9 | 22.8 | 13.7 |
| Age | Definitely | Probably | Probably Not | Definitely Not | Don't Know |
| Under 30 | 15.8 | 16.6 | 19.6 | 15.9 | 15.1 |
| 30-44 | 21.0 | 23.9 | 21.0 | 25.4 | 17.5 |
| 45-64 | 31.4 | 32.6 | 39.5 | 41.9 | 38.5 |
| 65 and older | 31.8 | 27.0 | 19.9 | 16.8 | 28.9 |
| Gender | Definitely | Probably | Probably Not | Definitely Not | Don't Know |
| Female | 42.7 | 47.7 | 56.0 | 60.9 | 62.1 |
| Male | 57.3 | 52.3 | 44.0 | 39.1 | 37.9 |
| Race/Ethnicity | Definitely | Probably | Probably Not | Definitely Not | Don't Know |
| White | 81.2 | 75.0 | 72.2 | 65.6 | 70.5 |
| Black | 4.9 | 9.1 | 13.1 | 18.9 | 17.2 |
| Hispanic | 7.6 | 9.7 | 8.9 | 10.8 | 9.2 |
| Asian | 4.4 | 4.5 | 3.4 | 2.0 | 1.6 |
| Other | 1.9 | 1.7 | 2.3 | 2.6 | 1.5 |
| Education | Definitely | Probably | Probably Not | Definitely Not | Don't Know |
| HS or less | 25.6 | 28.9 | 30.0 | 37.7 | 40.0 |
| College | 28.1 | 30.8 | 38.5 | 36.0 | 32.2 |
| College grad | 25.9 | 24.4 | 19.6 | 19.4 | 18.8 |
| Postgrad | 20.4 | 15.9 | 11.9 | 6.9 | 9.0 |
| Region | Definitely | Probably | Probably Not | Definitely Not | Don't Know |
| Northeast | 18.7 | 19.1 | 20.0 | 14.8 | 16.8 |
| Midwest | 24.2 | 24.6 | 17.4 | 23.7 | 23.2 |
| South | 30.9 | 34.9 | 42.2 | 42.2 | 34.7 |
| West | 26.1 | 21.3 | 20.5 | 19.2 | 25.3 |
| Party | Definitely | Probably | Probably Not | Definitely Not | Don't Know |
| Democrat | 39.1 | 37.7 | 39.8 | 27.4 | 40.3 |
| Republican | 31.3 | 28.9 | 28.3 | 36.8 | 22.2 |
| Independent | 25.6 | 30.0 | 27.8 | 30.0 | 30.5 |
| Other | 4.1 | 3.4 | 4.1 | 5.8 | 7.0 |

**Table SI3:** Vaccine Hesitancy Reasons and Selected Variables

| *Q2: Reasons for Vaccine Hesitancy* |  |  |  |  |  |  |  |  |  |  |
| --- | --- | --- | --- | --- | --- | --- | --- | --- | --- | --- |
|  | Side Effects |  | Expensive |  | Don't need it |  | Don't work |  | Skeptical |  |
|  | No | Yes | No | Yes | No | Yes | No | Yes | No | Yes |
| Total survey | 46.0 | 54.0 | 91.5 | 8.5 | 83.8 | 16.2 | 60.3 | 39.7 | 70.0 | 30.0 |
| Age | No | Yes | No | Yes | No | Yes | No | Yes | No | Yes |
| Under 30 | 13.9 | 20.7 | 16.3 | 31.6 | 16.3 | 23.9 | 16.5 | 19.2 | 16.5 | 20.1 |
| 30-44 | 22.3 | 24.4 | 23.1 | 27.2 | 21.1 | 35.5 | 24.6 | 21.7 | 22.3 | 26.1 |
| 45-64 | 44.5 | 37.7 | 41.8 | 30.3 | 42.3 | 33.2 | 41.7 | 39.5 | 41.3 | 39.7 |
| 65+ | 19.4 | 17.2 | 18.9 | 11 | 20.3 | 7.4 | 17.3 | 19.6 | 20 | 14.1 |
| Gender | No | Yes | No | Yes | No | Yes | No | Yes | No | Yes |
| Female | 59.3 | 58.2 | 59.5 | 50.1 | 61.3 | 45.2 | 55.2 | 64.1 | 57.6 | 61.4 |
| Male | 40.7 | 41.8 | 40.5 | 49.9 | 38.7 | 54.8 | 44.8 | 35.9 | 42.4 | 38.6 |
| Race/Ethnicity | No | Yes | No | Yes | No | Yes | No | Yes | No | Yes |
| White | 70.6 | 67 | 68.5 | 69.7 | 68.8 | 67.9 | 67 | 71.1 | 71 | 63.2 |
| Black | 15.6 | 16.8 | 16.9 | 9.8 | 17 | 12.7 | 16 | 16.7 | 14 | 21.6 |
| Hispanic | 8.8 | 11 | 9.6 | 13.7 | 9 | 15 | 11.5 | 7.7 | 9.9 | 10.2 |
| Asian | 2.2 | 3 | 2.6 | 3.6 | 2.7 | 2.1 | 3.2 | 1.8 | 2.8 | 2.2 |
| Other | 2.8 | 2.2 | 2.4 | 3.3 | 2.5 | 2.3 | 2.3 | 2.7 | 2.3 | 2.8 |
| Education | No | Yes | No | Yes | No | Yes | No | Yes | No | Yes |
| HS or Less | 39.7 | 29.5 | 33.9 | 37.2 | 33.5 | 37.7 | 35.1 | 32.7 | 30.6 | 42.5 |
| College | 35.4 | 38.6 | 37.5 | 33.4 | 37 | 37.8 | 37.4 | 36.9 | 39.3 | 32.1 |
| College Grad | 16.9 | 21.7 | 19.3 | 21.3 | 19.6 | 18.9 | 18.8 | 20.5 | 19.9 | 18.6 |
| Postgrad | 8 | 10.2 | 9.3 | 8 | 9.9 | 5.6 | 8.7 | 9.9 | 10.2 | 6.8 |
| Region | No | Yes | No | Yes | No | Yes | No | Yes | No | Yes |
| Northeast | 17.4 | 16.9 | 17.8 | 10.7 | 17.7 | 14.5 | 16.6 | 18 | 18.2 | 14.7 |
| Midwest | 22 | 19.9 | 20.9 | 21.1 | 19.9 | 26.2 | 20.8 | 21 | 21.2 | 20.2 |
| South | 39.8 | 44.2 | 41.8 | 46.8 | 42.7 | 39.5 | 43 | 41 | 40 | 47.3 |
| West | 20.8 | 18.9 | 19.6 | 21.3 | 19.8 | 19.8 | 19.5 | 20.1 | 20.6 | 17.8 |
| Partisanship | No | Yes | No | Yes | No | Yes | No | Yes | No | Yes |
| Democrat | 31.2 | 34.6 | 32.2 | 41.6 | 37 | 12.5 | 26.7 | 42.6 | 37.8 | 22 |
| Republican | 35.4 | 30.8 | 33.5 | 26.2 | 31.6 | 39.6 | 36.2 | 28 | 28.8 | 42.6 |
| Independent | 28.2 | 29.7 | 29.3 | 25.8 | 27.5 | 37 | 31.5 | 25.3 | 28.6 | 30 |
| Other | 5.2 | 4.9 | 4.9 | 6.4 | 3.9 | 11 | 5.6 | 4.1 | 4.9 | 5.4 |

##### D.2 Row Percentage Cross-tabulations

**Table SI4:** Vaccine Hesitancy and Selected Variables (Row Percentages)

D.2 Row Percentage Cross-tabulations Table SI4: Vaccine Hesitancy and Selected Variables (Row Percentages)
| *Q1: If a vaccine for COVID-19 were available today*<br>*would you get the vaccine?* |  |  |  |  |  |
| --- | --- | --- | --- | --- | --- |
|  | Definitely | Probably | Probably Not | Definitely Not | Don't Know |
| Total survey | 20.3 | 24.2 | 18.9 | 22.8 | 13.7 |
| Age | Definitely | Probably | Probably Not | Definitely Not | Don't Know |
| Under 30 | 19.3 | 21.8 | 12.5 | 24.2 | 22.3 |
| 30-44 | 19.1 | 26.1 | 10.8 | 26 | 17.9 |
| 45-64 | 17.4 | 26.1 | 14.4 | 21.6 | 20.5 |
| 65 and older | 26.3 | 15.6 | 16.2 | 26.6 | 15.3 |
| Gender | Definitely | Probably | Probably Not | Definitely Not | Don't Know |
| Female | 16.3 | 26.1 | 16 | 21.7 | 19.9 |
| Male | 24.9 | 19.1 | 11.1 | 27.1 | 17.8 |
| Race/Ethnicity | Definitely | Probably | Probably Not | Definitely Not | Don't Know |
| White | 22.6 | 20.5 | 13.3 | 24.9 | 18.8 |
| Black | 8.1 | 34.9 | 19.1 | 17.8 | 20.1 |
| Hispanic | 16.5 | 26.5 | 13.6 | 25.3 | 18.1 |
| Asian | 27.2 | 14 | 6.5 | 33 | 19.3 |
| Other | 18.5 | 29.1 | 10.4 | 20.5 | 21.6 |
| Education | Definitely | Probably | Probably Not | Definitely Not | Don't Know |
| HS or less | 16.3 | 26.9 | 17.2 | 21.9 | 17.8 |
| College | 17.2 | 24.8 | 13.4 | 22.5 | 22 |
| College grad | 24 | 20.3 | 11.8 | 27 | 17 |
| Postgrad | 31.7 | 12 | 9.4 | 29.6 | 17.3 |
| Region | Definitely | Probably | Probably Not | Definitely Not | Don't Know |
| Northeast | 21.2 | 18.9 | 12.9 | 25.9 | 21.1 |
| Midwest | 21.6 | 23.8 | 14 | 26.1 | 14.5 |
| South | 16.9 | 25.9 | 12.8 | 22.8 | 21.5 |
| West | 23.8 | 19.7 | 15.7 | 23.3 | 17.5 |
| Party | Definitely | Probably | Probably Not | Definitely Not | Don't Know |
| Democrat | 21.9 | 17.3 | 15.1 | 25.2 | 20.5 |
| Republican | 21.2 | 27.8 | 10.1 | 23.1 | 17.8 |
| Independent | 17.9 | 24 | 14.5 | 25.3 | 18.2 |
| Other | 17.6 | 25.8 | 20.4 | 18 | 18.3 |

**Table SI5:**
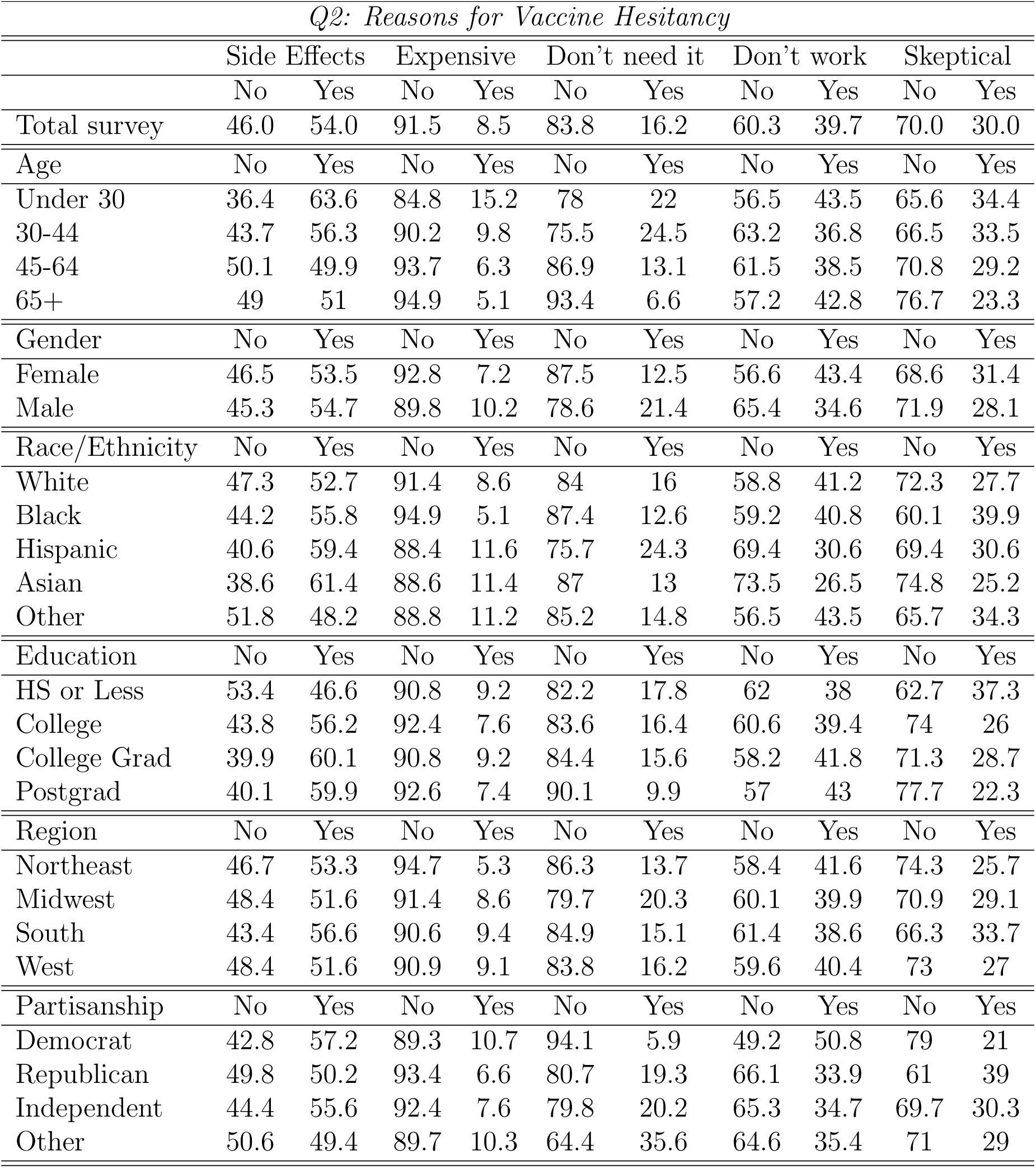
Vaccine Hesitancy Reasons and Selected Variables (Row Percentages)

#### E Multinomial Logit Regression

**Table SI6:** Multinomial Logit Regression (Point Estimates)

E Multinomial Logit Regression Table SI6: Multinomial Logit Regression (Point Estimates)
|  | COVID Vaccine Acceptance (Ref: Definitely Would) |  |  |  |
| --- | --- | --- | --- | --- |
|  | Probably Would<br>(1) | Probably Would Not<br>(2) | Definitely Would Not<br>(3) | Don't Know<br>(4) |
| **Age:**<br>(Ref: Under 30) |  |  |  |  |
| 30-44 | 0.117 (0.079) | 0.142 (0.089) | 0.079 (0.096) | 0.512*** (0.111) |
| 45-64 | 0.247*** (0.076) | 0.387*** (0.086) | 0.406*** (0.091) | 0.931*** (0.106) |
| 65+ | 0.204** (0.082) | 0.236** (0.095) | 0.087 (0.102) | 0.899*** (0.113) |
| **Gender:**<br>(Ref: Male) |  |  |  |  |
| Female | 0.164*** (0.044) | 0.708*** (0.052) | 1.024*** (0.055) | 0.593*** (0.056) |
| **Race:**<br>(Ref: White) |  |  |  |  |
| Black | 0.484*** (0.105) | 0.653*** (0.117) | 1.928*** (0.107) | 0.921*** (0.115) |
| Hispanic | -0.043 (0.065) | 0.352*** (0.072) | 0.495*** (0.077) | -0.023 (0.083) |
| Asian | 0.007 (0.067) | -0.004 (0.079) | -0.183** (0.092) | -0.266*** (0.091) |
| Other | -0.110 (0.096) | 0.430*** (0.100) | 0.741*** (0.100) | 0.594*** (0.104) |
| **Education:**<br>(Ref: HS or Less) |  |  |  |  |
| Some College | 0.211** (0.099) | 0.175* (0.105) | -0.314*** (0.102) | -0.256** (0.107) |
| College Grad | 0.156* (0.088) | 0.019 (0.093) | -0.622*** (0.089) | -0.532*** (0.094) |
| Postgrad | -0.003 (0.103) | -0.324*** (0.112) | -0.758*** (0.109) | -0.442*** (0.111) |
| **Census Region:**<br>(Ref: Northeast) |  |  |  |  |
| Midwest | 0.037 (0.217) | -0.368 (0.244) | -0.037 (0.262) | 0.458* (0.271) |
| South | -0.124 (0.199) | -0.085 (0.216) | 0.240 (0.234) | 0.068 (0.259) |
| West | -0.183 (0.162) | -0.505*** (0.178) | -0.226 (0.199) | 0.032 (0.218) |
| **Political News:**<br>(Ref: Balanced) |  |  |  |  |
| Right-wing | -0.321*** (0.075) | -0.728*** (0.089) | -0.852*** (0.086) | -0.481*** (0.100) |
| Left-wing | -0.074 (0.052) | -0.166*** (0.059) | -0.183*** (0.063) | 0.029 (0.064) |
| **COVID-19 Spread:**<br>(Ref: Not Worried) |  |  |  |  |
| Very Worried | -0.663*** (0.122) | -1.265*** (0.134) | -2.128*** (0.123) | -0.671*** (0.154) |
| Somewhat Worried | -0.303** (0.118) | -0.965*** (0.128) | -1.862*** (0.117) | -0.338** (0.150) |
| Not too Worried | -0.379*** (0.117) | -0.439*** (0.124) | -1.079*** (0.111) | -0.218 (0.147) |
| **Loss of Income** | 0.151*** (0.049) | 0.036 (0.059) | 0.071 (0.060) | -0.035 (0.061) |
| **Financial Situation:**<br>(Ref: Not Worried) |  |  |  |  |
| Very Worried | 0.019 (0.085) | 0.436*** (0.100) | 0.599*** (0.095) | 0.248** (0.105) |
| Somewhat Worried | 0.755*** (0.067) | 1.042*** (0.081) | 0.716*** (0.081) | 0.740*** (0.085) |
| Not too Worried | 0.495*** (0.061) | 0.590*** (0.078) | 0.352*** (0.076) | 0.610*** (0.077) |
| **Poor in COVID:**<br>(Yes/No) |  |  |  |  |
| Trump | -0.114 (0.108) | -0.329*** (0.120) | 0.265** (0.128) | 0.172 (0.134) |
| CDC | 0.229*** (0.075) | 0.593*** (0.079) | 1.197*** (0.075) | 0.248*** (0.093) |
| Local Gov. | -0.096 (0.072) | 0.201*** (0.076) | 0.254*** (0.074) | -0.054 (0.092) |
| US Gov. | 0.169** (0.085) | 0.359*** (0.094) | 0.498*** (0.098) | 0.366*** (0.104) |
| State Gov. | -0.139** (0.070) | -0.157** (0.079) | 0.078 (0.079) | -0.369*** (0.089) |
| **Reopen too Quick** | 0.071 (0.061) | 0.035 (0.066) | -0.259*** (0.077) | 0.092 (0.079) |
| **Social Issues:**<br>(Important/Not) |  |  |  |  |
| Immigration | 0.031 (0.061) | -0.005 (0.069) | 0.138* (0.076) | 0.0003 (0.075) |
| Abortion | 0.340*** (0.052) | 0.093 (0.059) | 0.262*** (0.063) | -0.243*** (0.062) |
| Foreign Policy | 0.237*** (0.065) | 0.341*** (0.077) | 0.238*** (0.083) | 0.524*** (0.086) |
| Inequality | 0.166** (0.074) | 0.114 (0.088) | 0.006 (0.093) | -0.028 (0.098) |
| COVID-19 | -0.146* (0.088) | -0.429*** (0.101) | -0.581*** (0.093) | -0.128 (0.108) |
| Crime | 0.039 (0.058) | 0.246*** (0.068) | 0.455*** (0.073) | 0.132* (0.070) |
| Economy | -0.429*** (0.084) | -0.329*** (0.099) | -0.293** (0.118) | -0.293*** (0.112) |
| Healthcare | -0.669*** (0.096) | -0.323*** (0.110) | -0.309*** (0.115) | -0.377*** (0.127) |
| Race Equality | 0.180** (0.078) | 0.233*** (0.087) | 0.156* (0.089) | 0.505*** (0.098) |
| Climate | -0.116 (0.078) | 0.185** (0.088) | -0.021 (0.090) | -0.370*** (0.093) |
| Guns | -0.138** (0.054) | -0.229*** (0.063) | -0.075 (0.071) | 0.013 (0.068) |
| SCOTUS | 0.011 (0.069) | -0.248*** (0.075) | -0.374*** (0.079) | 0.082 (0.087) |
| **Democratic** | -0.145** (0.057) | -0.395*** (0.064) | -0.286*** (0.072) | -0.153* (0.078) |
| **Republican** | 0.284*** (0.076) | 0.023 (0.091) | 0.377*** (0.097) | -0.468*** (0.101) |
| **Support BLM** | 0.177** (0.075) | -0.093 (0.085) | -0.723*** (0.087) | -0.119 (0.094) |
| **Follow Public Affairs** | -0.172** (0.075) | -0.533*** (0.078) | -0.256*** (0.084) | -0.830*** (0.083) |
| **Income:**<br>(Ref: Under 50K) |  |  |  |  |
| 50K-100K | 0.179** (0.081) | -0.225** (0.090) | -0.045 (0.088) | -0.121 (0.086) |
| 100K+ | 0.107 (0.099) | -0.102 (0.098) | -0.140 (0.087) | -0.277*** (0.098) |
| **Social Media Use:**<br>(Ref: Infrequent) |  |  |  |  |
| Light | 0.245*** (0.059) | 0.332*** (0.071) | 0.552*** (0.077) | 0.186*** (0.071) |
| Heavy | 0.686*** (0.096) | 0.661*** (0.113) | 0.662*** (0.125) | 0.347*** (0.128) |
| Akaike Inf. Crit. | 54,727.890 | 54,727.890 | 54,727.890 | 54,727.890 |

Note:
\*p<0.1; \*\*p<0.05; \*\*\*p<0.01

#### F Binary Logit Regression

**Table SI7:** Binary Logit Regression (Average Marginal Effects) (Part 1)

F Binary Logit Regression Table SI7: Binary Logit Regression (Average Marginal Effects) (Part 1)
|  | COVID Vaccine Acceptance |  |  |
| --- | --- | --- | --- |
|  | Acceptance/Refusal<br>(1) | Refusal:Side Effect<br>(2) | Refusal:Too Expensive<br>(3) |
| **Age:**<br>(Ref: Under 30) |  |  |  |
| 30-44 | -0.031 (0.035) | -0.098** (0.047) | -0.041 (0.029) |
| 45-64 | -0.085*** (0.032) | -0.194*** (0.044) | -0.076*** (0.028) |
| 65+ | 0.042 (0.038) | -0.212*** (0.058) | -0.082** (0.034) |
| **Gender:**<br>(Ref: Male) |  |  |  |
| Female | -0.145*** (0.021) | -0.013 (0.034) | -0.046** (0.019) |
| **Race:**<br>(Ref: White) |  |  |  |
| Black | -0.186*** (0.037) | -0.010 (0.048) | -0.070*** (0.017) |
| Hispanic | -0.035 (0.037) | 0.046 (0.050) | -0.009 (0.026) |
| Asian | 0.086 (0.060) | 0.059 (0.112) | 0.015 (0.051) |
| Other | -0.065 (0.049) | -0.060 (0.072) | -0.008 (0.042) |
| **Education:**<br>(Ref: HS or Less) |  |  |  |
| Some College | -0.023 (0.029) | 0.091** (0.039) | -0.015 (0.020) |
| College Grad | 0.071** (0.028) | 0.113*** (0.041) | 0.008 (0.022) |
| Postgrad | 0.107*** (0.036) | 0.101* (0.059) | -0.012 (0.030) |
| **Census Region:**<br>(Ref: Northeast) |  |  |  |
| Midwest | 0.044 (0.034) | 0.006 (0.052) | 0.035 (0.024) |
| South | -0.038 (0.032) | 0.034 (0.046) | 0.052*** (0.020) |
| West | 0.038 (0.030) | -0.025 (0.047) | 0.041** (0.020) |
| **Political News:**<br>(Ref: Balanced) |  |  |  |
| Right-wing | 0.103*** (0.030) | -0.005 (0.045) | -0.017 (0.022) |
| Left-wing | -0.013 (0.025) | 0.015 (0.039) | -0.013 (0.020) |
| **COVID-19 Spread:**<br>(Ref: Not Worried) |  |  |  |
| Very Worried | 0.251*** (0.047) | 0.314*** (0.061) | 0.016 (0.031) |
| Somewhat Worried | 0.228*** (0.045) | 0.251*** (0.057) | 0.027 (0.032) |
| Not too Worried | 0.149*** (0.040) | 0.138*** (0.052) | 0.062** (0.031) |
| **Loss of Income** | -0.040* (0.022) | 0.023 (0.033) | 0.030* (0.018) |
| **Financial Situation:**<br>(Ref: Not Worried) |  |  |  |
| Very Worried | -0.029 (0.039) | -0.045 (0.061) | 0.062* (0.033) |
| Somewhat Worried | -0.037 (0.033) | 0.014 (0.051) | 0.050** (0.025) |
| Not too Worried | -0.014 (0.032) | 0.011 (0.049) | 0.025 (0.022) |
| **Poor in COVID:**<br>(Yes/No) |  |  |  |
| Trump | 0.036 (0.047) | -0.035 (0.069) | 0.009 (0.030) |
| CDC | -0.104*** (0.033) | 0.071 (0.043) | 0.028 (0.026) |
| Local Gov. | -0.056** (0.028) | 0.030 (0.040) | -0.023 (0.019) |
| US Gov. | -0.120*** (0.033) | -0.122** (0.048) | -0.046 (0.029) |
| State Gov. | 0.005 (0.027) | 0.016 (0.040) | -0.025 (0.025) |
| **Reopen too Quick** | 0.045 (0.030) | -0.070* (0.042) | 0.035 (0.023) |
| **Social Issues:**<br>(Important/Not) |  |  |  |
| Immigration | 0.015 (0.030) | -0.008 (0.046) | 0.020 (0.021) |
| Abortion | 0.024 (0.024) | 0.006 (0.037) | -0.005 (0.020) |
| Foreign Policy | 0.045 (0.032) | 0.003 (0.047) | 0.007 (0.024) |
| Inequality | -0.071** (0.034) | -0.084* (0.050) | 0.033 (0.028) |
| COVID-19 | 0.110*** (0.034) | 0.027 (0.051) | -0.026 (0.031) |
| Crime | -0.039 (0.029) | 0.011 (0.048) | 0.023 (0.021) |
| Economy | -0.013 (0.041) | -0.046 (0.065) | 0.017 (0.029) |
| Healthcare | -0.027 (0.037) | 0.166*** (0.057) | 0.029 (0.024) |
| Race Equality | 0.012 (0.034) | 0.072 (0.051) | -0.050 (0.035) |
| Climate | 0.032 (0.035) | -0.051 (0.049) | 0.017 (0.026) |
| Guns | -0.017 (0.026) | 0.066 (0.045) | 0.005 (0.020) |
| SCOTUS | -0.029 (0.031) | -0.095** (0.045) | -0.063* (0.034) |
| **Democratic** | 0.047* (0.028) | -0.020 (0.045) | 0.023 (0.023) |
| **Republican** | 0.015 (0.031) | -0.019 (0.044) | 0.010 (0.025) |
| **Support BLM** | 0.065* (0.036) | -0.003 (0.048) | 0.028 (0.027) |
| **Follow Public Affairs** | 0.047 (0.033) | 0.096** (0.044) | 0.009 (0.024) |
| **Income:**<br>(Ref: Under 50K) |  |  |  |
| 50K-100K | 0.015 (0.027) | 0.026 (0.040) | 0.017 (0.022) |
| 100K+ | 0.043 (0.029) | 0.041 (0.051) | -0.025 (0.022) |
| **Social Media Use:**<br>(Ref: Infrequent) |  |  |  |
| Light | -0.011 (0.031) | -0.017 (0.049) | -0.018 (0.038) |
| Heavy | -0.032 (0.048) | -0.080 (0.073) | 0.005 (0.046) |
| Observations | 4,362 | 1,897 | 1,897 |
| AUC-ROC | 0.7214 | 0.6291 | 0.7130 |

Note:
\*p<0.1; \*\*p<0.05; \*\*\*p<0.01

**Table SI8:** Binary Logit Regression (Average Marginal Effects) (Part 2)

|  | COVID Vaccine Acceptance |  |  |
| --- | --- | --- | --- |
|  | Refusal:Don't Need<br>(1) | Refusal:Don't Know<br>(2) | Refusal:Skeptical<br>(3) |
| **Age:**<br>(Ref: Under 30) |  |  |  |
| 30-44 | 0.008 (0.034) | -0.069 (0.050) | -0.019 (0.049) |
| 45-64 | -0.090** (0.035) | -0.064 (0.047) | -0.054 (0.047) |
| 65+ | -0.129*** (0.040) | -0.014 (0.059) | -0.113** (0.056) |
| **Gender:**<br>(Ref: Male) |  |  |  |
| Female | -0.008 (0.020) | 0.021 (0.032) | 0.075** (0.032) |
| **Race:**<br>(Ref: White) |  |  |  |
| Black | 0.057 (0.040) | -0.067 (0.045) | 0.187*** (0.050) |
| Hispanic | 0.066 (0.040) | -0.131*** (0.047) | 0.046 (0.048) |
| Asian | -0.049 (0.043) | -0.149** (0.071) | 0.031 (0.101) |
| Other | -0.008 (0.035) | -0.002 (0.074) | 0.091 (0.069) |
| **Education:**<br>(Ref: HS or Less) |  |  |  |
| Some College | -0.016 (0.023) | 0.008 (0.039) | -0.088** (0.035) |
| College Grad | 0.009 (0.026) | 0.033 (0.039) | -0.048 (0.038) |
| Postgrad | 0.0004 (0.040) | 0.025 (0.057) | -0.049 (0.055) |
| **Census Region:**<br>(Ref: Northeast) |  |  |  |
| Midwest | 0.032 (0.031) | 0.024 (0.051) | 0.005 (0.049) |
| South | 0.007 (0.028) | 0.014 (0.045) | 0.055 (0.043) |
| West | 0.002 (0.026) | 0.034 (0.044) | 0.009 (0.044) |
| **Political News:**<br>(Ref: Balanced) |  |  |  |
| Right-wing | -0.017 (0.021) | -0.035 (0.046) | 0.082** (0.042) |
| Left-wing | -0.101*** (0.025) | -0.007 (0.037) | 0.029 (0.038) |
| **COVID-19 Spread:**<br>(Ref: Not Worried) |  |  |  |
| Very Worried | -0.279*** (0.042) | 0.073 (0.064) | 0.038 (0.052) |
| Somewhat Worried | -0.216*** (0.044) | 0.152** (0.060) | 0.120*** (0.045) |
| Not too Worried | -0.109*** (0.038) | 0.110** (0.055) | 0.075* (0.042) |
| **Loss of Income** | 0.067*** (0.023) | 0.019 (0.033) | -0.027 (0.031) |
| **Financial Situation:**<br>(Ref: Not Worried) |  |  |  |
| Very Worried | -0.043 (0.031) | 0.142** (0.058) | 0.017 (0.056) |
| Somewhat Worried | -0.014 (0.029) | 0.077 (0.049) | -0.036 (0.044) |
| Not too Worried | 0.021 (0.027) | 0.088* (0.047) | -0.036 (0.043) |
| **Poor in COVID:**<br>(Yes/No) |  |  |  |
| Trump | -0.007 (0.052) | 0.023 (0.068) | 0.021 (0.069) |
| CDC | 0.019 (0.024) | -0.068* (0.041) | 0.077* (0.040) |
| Local Gov. | 0.058** (0.025) | 0.005 (0.041) | -0.002 (0.037) |
| US Gov. | -0.060** (0.028) | 0.038 (0.050) | 0.018 (0.046) |
| State Gov. | -0.003 (0.023) | 0.015 (0.040) | 0.074** (0.036) |
| **Reopen too Quick** | -0.015 (0.029) | -0.032 (0.039) | -0.072* (0.039) |
| **Social Issues:**<br>(Important/Not) |  |  |  |
| Immigration | 0.033 (0.028) | -0.071* (0.041) | -0.008 (0.047) |
| Abortion | -0.023 (0.023) | 0.022 (0.036) | 0.019 (0.034) |
| Foreign Policy | 0.0004 (0.028) | -0.003 (0.045) | -0.018 (0.045) |
| Inequality | 0.003 (0.029) | 0.051 (0.051) | -0.018 (0.045) |
| COVID-19 | 0.015 (0.026) | 0.082* (0.048) | -0.119** (0.048) |
| Crime | -0.002 (0.030) | 0.013 (0.043) | 0.030 (0.047) |
| Economy | 0.021 (0.039) | 0.069 (0.062) | 0.080 (0.060) |
| Healthcare | -0.093*** (0.032) | -0.002 (0.056) | -0.056 (0.049) |
| Race Equality | -0.005 (0.027) | -0.067 (0.049) | 0.061 (0.044) |
| Climate | 0.022 (0.029) | -0.020 (0.048) | -0.004 (0.046) |
| Guns | -0.011 (0.026) | 0.033 (0.041) | -0.079* (0.046) |
| SCOTUS | -0.034 (0.030) | -0.057 (0.045) | 0.017 (0.044) |
| **Democratic** | -0.021 (0.041) | 0.060 (0.046) | -0.004 (0.049) |
| **Republican** | -0.060*** (0.023) | 0.039 (0.044) | 0.031 (0.038) |
| **Support BLM** | -0.044 (0.034) | 0.038 (0.047) | -0.038 (0.045) |
| **Follow Public Affairs** | 0.029 (0.024) | -0.004 (0.042) | 0.021 (0.038) |
| **Income:**<br>(Ref: Under 50K) |  |  |  |
| 50K-100K | -0.019 (0.023) | 0.011 (0.037) | -0.003 (0.035) |
| 100K+ | -0.045 (0.027) | 0.035 (0.045) | -0.047 (0.043) |
| **Social Media Use:**<br>(Ref: Infrequent) |  |  |  |
| Light | 0.022 (0.029) | 0.076 (0.047) | -0.080 (0.049) |
| Heavy | 0.080* (0.049) | 0.023 (0.069) | -0.075 (0.068) |
| Observations | 1,897 | 1,897 | 1,897 |
| AUC-ROC | 0.8550 | 0.6554 | 0.7010 |

Note:
\*p<0.1; \*\*p<0.05; \*\*\*p<0.01

#### G Complete Plots of Average Marginal Effects

**Figure SI2:**
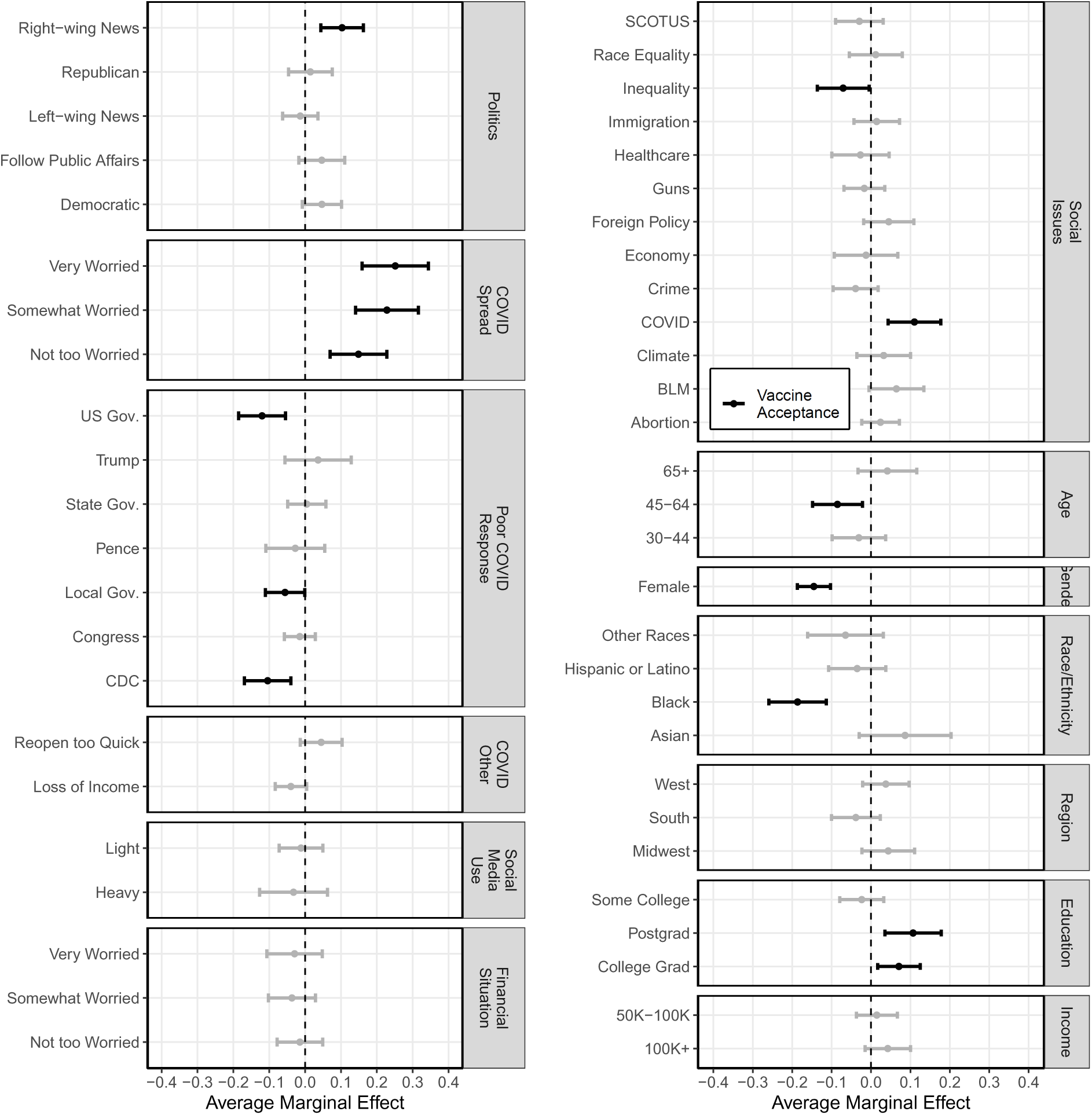
Logit Regression: Vaccine Acceptance (Complete)

**Figure SI3:**
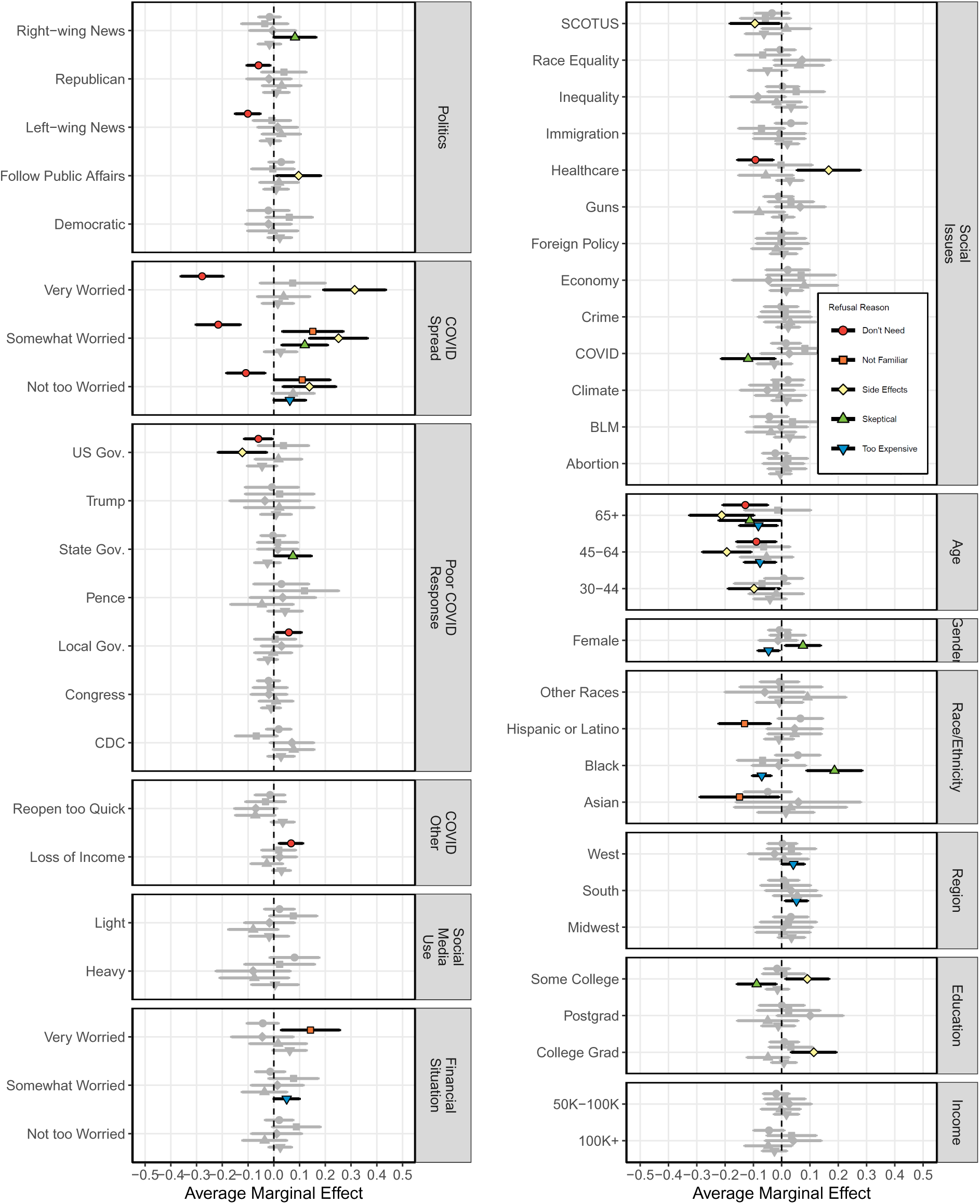
Logit Regression: Refusal Reasons (Complete)

## Footnotes

1 https://www.fda.gov/regulatory-information/search-fda-guidance-documents/emergency-use-authorization-vaccines-prevent-covid-19.

2 https://covid.cdc.gov/covid-data-tracker/#vaccinations, last accessed November 4, 2021.

3 https://bit.ly/35b2BiJ, last accessed June 10, 2021.

4 https://www.pfizer.com/news/press-release/press-release-detail/pfizer-and-biontech-announce-vaccine-candidate-against, last accessed May 28, 2021

5 https://www.who.int/news-room/spotlight/ten-threats-to-global-health-in-2019

6 KAP COVID Trend Analysis, see https://bit.ly/3cA1tt8, last accessed June 10, 2021

7 Readers interested in seeing cross-tabulations of this question against a number of other survey questions will find those results in the Supporting Information.

8 Specifically, respondents who said in response to Q1 that they “probably would not” or ”definitely would not” get a vaccine were it available to them were asked these follow-up questions; respondents who indicated they might or would get the vaccine, or who did not answer Q1, did not receive Q2.

9 Cross-tabulations of these responses against other independent variables are available in the paper’s Supporting Information.

10 Figures with the complete average marginal effects results are provided in the Supporting Information, Section G.

11 Complete results from the logistic models are presented in the Supporting Information, Section F.

12 The respondents were asked to select one or more news sources that they use frequently. The right-wing sources include Fox, WSJ, NYP, Washington Examiner, Breibart, and The Drudge Report. The left-wing sources include CNN, NBC, NYT, Washington Post, HuffPost, and Yahoo!. The mixed sources include USA Today and Google. We define voters who follow more right-wing news sources than the left-wing news sources as right-wing news followers

13 The respondents were asked to select one or more social networks that they use every day. The social networks include Twitter, Facebook, Instagram, WhatsApp, YouTube, WeChat, Snapchat, Linkedin, and TikTok. We define respondents who use 1-4 social networks daily as light users, 5-9 as heavy users, and 0 as infrequent users.

